# Genetic Determinants of Bone Microarchitecture and its Association with Health Outcomes: A Genome-wide Association and Mendelian Randomization Study on Trabecular Bone Score

**DOI:** 10.1101/2024.10.06.24314958

**Authors:** Sijia Guo, Jiping Zhang, Huiwu Li, Guan Ning Lin, Cheng-Kung Cheng, Jingwei Zhang

## Abstract

**Background:** Bone microarchitecture is a critical determinant of bone strength and fracture risk, yet its genetic basis and relationship to systemic health remain largely unexplored. This study aimed to identify genetic determinants of bone microarchitecture using trabecular bone score (TBS) and investigate the causal relationships between bone microarchitecture and various health outcomes.

**Methods:** We conducted a genome-wide association study (GWAS) of TBS in 25,268 UK Biobank participants to identify genetic loci associated with bone microarchitecture. Two-sample Mendelian randomization (MR) was employed to assess the causal relationships between systemic health risk factors and bone microarchitecture, as well as the impact of bone microarchitecture on musculoskeletal disorders.

**Findings:** The GWAS identified 75 significant single nucleotide polymorphisms (SNPs) across 19 genomic loci, with an estimated heritability of TBS at 24.5%. Many of these loci (18/19) were also associated with bone mineral density (BMD) and fractures, indicating a shared genetic basis for bone microarchitecture and bone mass. MR analysis revealed that rheumatoid arthritis has a significant causal effect on the deterioration of bone microarchitecture (β = -0.003, P = 1.14×10^-4^). Suggestive associations were found between bone microarchitecture deterioration and inflammatory bowel disease, cardiovascular disease, and depression (P < 0.05). Moreover, genetically predicted TBS was significantly associated with fracture risk (OR = 0.003, P = 1.89×10^-8^) and suggestively associated with osteonecrosis (OR = 0.002, P = 0.040).

**Interpretation:** This study identified novel genetic determinants of bone microarchitecture and demonstrated its association with various systemic diseases, highlighting the critical role of bone microarchitecture in skeletal health. The results advocate for the clinical use of TBS to better assess the risk of osteoporosis and fractures and to improve bone and overall health assessments. The causal effect of rheumatoid arthritis on microarchitectural deterioration underscores the need for increased monitoring of bone health in this population.

**Funding:** This work supported by Shanghai "Rising Stars of Medical Talent" Youth Development Program, Youth Medical Talents-Specialist Program (grant number SHHWRS 2023-62), the Fundamental Research Funds for the Central Universities (grant number AF0820060), Outstanding Research-oriented Doctor Cultivation Program at the Ninth People’s Hospital affiliated with the School of Medicine, Shanghai Jiao Tong University, National Natural Science Foundation of China (grant number 31900941).

## 1. Introduction

Osteoporosis (OP) is a multifactorial bone disease characterized by decreased bone mass and deteriorated bone microarchitecture, leading to lower bone strength and increased susceptibility to fractures^1,2^. OP results from genetic and environmental factors and their complex interactions^3,4^. OP is a significant public health problem, and the healthcare and socioeconomic impacts of osteoporotic fractures are expected to increase as the global population ages^5,6^.

Bone mineral density (BMD) is the cornerstone for the diagnosis of OP and assessment of fracture risk^7^. Consequently, numerous genetic loci associated with BMD and fracture risk have been identified^8–10^. However, the genetic factors influencing bone microarchitecture, another crucial determinant of bone quality and strength^11,12^, remain largely unknown. The trabecular bone score (TBS), derived from dual-energy X-ray absorptiometry (DXA) images, is a promising surrogate measure of trabecular microstructure and vertebral fracture risk^13,14^. In vivo bone biopsy studies have shown a significant correlation between TBS and bone microstructural parameters^15^, suggesting that TBS is a promising index for investigating the genetic basis of bone microarchitecture. A heritability analysis using a biometric model on data from 265 families estimated that approximately 45% of the variance in TBS is attributable to genetic factors^16^. A modest-scale genome-wide association study (GWAS)^17^ identified two intronic variants—rs1815994 and rs11630730—were significantly associated with TBS in community-dwelling adults older than 50 years. However, these findings only represent a small fraction of the complex genetic landscape contributing to bone microarchitectural deterioration. Therefore, a comprehensive large-scale investigation is necessary to further elucidate the genetic basis of bone microarchitecture.

Observational studies have identified risk factors for bone microarchitecture deterioration and its association with musculoskeletal disorders^18–21^. Nonetheless, the causal relationships between these risk factors and disorders are unclear. Mendelian randomization (MR)^22^ is a powerful statistical method that uses genetic variants as instrumental variables (IVs) to reduce confounding and reverse causation, thus more accurately estimating the causal relationship between exposure and outcome. Genetic variants are inherited from parents and are randomly distributed; therefore, these variants are unaffected by environmental and other confounders. Large-scale GWAS of bone microarchitecture can improve MR analyses and accurately estimate causal effects.

This study evaluated (1) the effect of genetic factors on bone microarchitecture by performing a GWAS of UK Biobank data and (2) causal associations between TBS, clinical risk factors, and musculoskeletal disorders by MR analysis. The methodological approach used in this study can elucidate the complexity of bone microarchitecture and its role in health outcomes and can help guide interventions to improve bone health.

## 2. Methods

### 2.1 Cohort and genetic information

Over 500,000 participants aged 40-69 years were enrolled in the UK Biobank between 2006 and 2010 and are being followed up. All participants provided informed consent to use their health records for research purposes. This study was approved by the National Health Service Research Ethics Committee (Approval No. 11/NW/0382). DNA extraction, quality control (QC), and genotyping were performed using standardized protocols, and the details can be found on the UK Biobank website (https://www.ukbiobank.ac.uk/). Whole-genome sequencing data of 501,708 samples were released in 2017, and the QC, imputation, and analysis of these data were performed as described previously^23^. We applied for and received access to phenotypic and genotypic data from the UK Biobank (Grant Number: 93966).

### 2.2 Phenotypic information and study population

TBSs from the UK Biobank served as a surrogate phenotype to identify genetic loci associated with bone microarchitecture. TBS is calculated based on experimental variograms of 2D projected DXA gray-level images^14^. DXA-based TBSs of the lumbar spine (segments L1 to L4) were calculated as described in a previous study^24^ and at https://biobank.ndph.ox.ac.uk/showcase/field.cgi?id=21005.

### 2.3 Statistical analysis

#### 2.3.1 GWAS analysis

GWAS was performed using REGENIE^25^ (version 2.2), which performs regression analyses of large genome datasets. The analyses were carried out in two steps. In the first step, sparse genotype calls were used to fit a genome-wide regression model to the TBS, generating a set of genomic predictions as output. SNPs with INFO scores < 0.1, minor allele frequencies (MAF) < 0.01, and P < 1×10^-15^ in the Hardy-Weinberg equilibrium (HWE) test were excluded. Principal component analysis was performed to identify and correct for population structure in the samples. Data from individuals of non-White British ancestry, individuals related to at least one other participant in the cohort (with a relatedness threshold of 0.025), and individuals who did not pass QC were excluded. Then, age, gender, BMI, and ten population principal components were adjusted as covariates in the regression. In the second step, association tests were performed on the SNPs of densely imputed genotypes based on logistic regression with Firth correction and the QC method described above.

After these two steps, quantitative associations between 509,485 variants and TBSs from 25,268 individuals were analyzed. In GWAS analyses, genome-wide significance was set at P < 5 × 10^-8^; independent SNPs were defined as those not associated with other relevant SNPs (R^2^ < 0.6), and linkage disequilibrium (LD) blocks were identified for independent SNPs in the 250-kb range.

#### 2.3.2 Post-GWAS analysis

Functional mapping and annotation were performed using FUMA^26^, and gene-based association analysis was performed using MAGMA version 1.6^27^, which maps SNPs to protein-coding genes from Ensembl build 85^28^. Default parameters and the Molecular Signatures Database version 7.0^29^ were used in gene set analysis. Tissue-level expression was analyzed using GTEx data^30^ and MAGMA. P-values were adjusted using Bonferroni correction.

#### 2.3.3 Genetic correlation analysis

Genetic correlations between bone microarchitecture and 48 complex traits were estimated using LD score regression^31^. These traits are related to diseases, anthropometric parameters, lifestyle factors, hematological indicators, and social factors. The genetic determinants of these traits were derived from large-scale GWAS from the Open GWAS Project supported by the Integrative Epidemiology Unit (https://gwas.mrcieu.ac.uk/)^32^. After strict Bonferroni correction, the significant threshold should be 0.001 (0.05/48). P-values between 0.001 and 0.05 were considered suggestively significant.

#### 2.3.4 MR analysis

The causal relationship between bone microarchitecture and health outcomes was assessed using two-sample MR. Using this approach, we assessed the causal effects of 17 risk factors on bone microarchitecture and the causal effects of bone microarchitecture on musculoskeletal disorders. The genetic determinants of these risk factors were derived from the largest publicly available GWAS dataset containing data on age of smoking initiation^33^, number of cigarettes per day^33^, alcohol consumption^34^, number of alcoholic drinks per week^33^, age at menopause^35^, age at menarche^36^, vitamin D levels^37^, rheumatoid arthritis (RA)^37^, inflammatory bowel disease^38^, grip strength^39^, type 2 diabetes^40^, fasting glucose^41^, coronary heart disease^42^, cardiovascular diseases^43^, fibroblastic disorders^37^, asthma^37^, and major depression^44^. Data on the genetic determinants of musculoskeletal disorders, including osteoporosis, fractures, osteonecrosis, ankylosing spondylitis, low back pain, spinal stenosis, and RA, were collected from FinnGen^37^, and data on the genetic determinants of knee and hip osteoarthritis were obtained from arcOGEN^45^.

Causal effects were estimated using inverse variance weighting. IVs were screened to select those strongly correlated with exposure and those not affected by confounders and to ensure that there was no genetic pleiotropy, i.e., the effects of IVs on the outcome were mediated solely by the exposure^46^. The strength of SNPs was evaluated using F statistics, and IVs with F < 10 and P > 5 × 10^-8^ were excluded to avoid weak instrument bias. IVs associated with potential confounders were excluded using PhenoScanner version 2 (http://www.phenoscanner.medschl.cam.ac.uk/)^47^. SNPs were clustered using a stringent criterion, and those with high LD were excluded to ensure SNP independence.

Four sensitivity analyses—weighted median^48^, MR-Egger regression^49^, Mendelian randomization pleiotropy residual sum and outlier (MR-PRESSO)^50^, and leave-one-out approach—were performed to test the reliability of MR findings and avoid false positives due to potential pleiotropic effects of genetic variants. Leave-one-out analysis was performed to evaluate the influence of potentially pleiotropic SNPs on causal estimates^51^. Bonferroni-corrected P-values of 0.002 and 0.004 (0.05/17 risk factors and 0.05/12 musculoskeletal disorders, respectively) were used to identify significant correlations, and P-values between 0.002 and 0.05 and between 0.004 and 0.05 were considered suggestively significant. All statistical analyses were performed using the “TwoSampleMR” and “MRPRESSO” packages in R version 4.0.5.

## 3. Results

### 3.1 Baseline characteristics

After screening, 25,268 UK Biobank participants with data on TBS, complete genotypes, and covariates were included in the study. The baseline characteristics of the participants (N = 25,268) are shown in Table 1. The mean age and body mass index (BMI) of the cohort was 56 years and 26.1 kg/m2, and 51.46% were female.

**Table 1.** Baseline characteristics.

| **Characteristic** | **Overall, N = 25,268<sup>1</sup>** | **Female, N = 13,002<sup>1</sup>** | **Male, N = 12,266<sup>1</sup>** |
| --- | --- | --- | --- |
| **Age** | 56 (49, 61) | 55 (49, 60) | 57 (50, 62) |
| **Body mass index** | 26.1 (23.7, 28.9) | 25.3 (23.0, 28.5) | 26.8 (24.6, 29.2) |
| **Trabecular bone score** | 1.15 (1.09, 1.21) | 1.13 (1.07, 1.19) | 1.17 (1.12, 1.23) |
| **Spine BMD** | 1.09 (0.96, 1.21) | 0.99 (0.90, 1.10) | 1.18 (1.09, 1.29) |
| **Femoral neck BMD** | 0.93 (0.84, 1.04) | 0.89 (0.80, 0.98) | 0.98 (0.90, 1.08) |
| **Any fracture\*** | 1,969 (7.8%) | 1,057 (8.2%) | 912 (7.5%) |

<sup>1</sup> Median (IQR); n (%)
BMD, bone mineral density. \*fractured or broken bones in the last 5 years.

### 3.2 GWAS of TBS

GWAS of TBS values of 25,268 white British individuals identified 75 independent significant SNPs across 19 genetic loci (P < 5 × 10^-8^; Fig. 2; Tables S2-S4). All significant candidate SNPs (N = 2,217) are shown in Table S5. The most significant risk locus for microarchitectural deterioration was located in chromosome 1p31.3, which contains the Wntless (*WLS*) gene, with a P-value of 2.14×10^-32^ for the lead SNPs rs7062, rs2566752, and rs7554551. The second most significant locus was located in the 13q14.11 region, with a P-value of 7.68×10^-25^ for the lead SNPs 13:42949052_TC_T, rs548589580, and rs78667121.

The Q–Q plot of GWAS is shown in Fig. 1. LD score regression analysis showed that the heritability of TBS was 24.52% (SEM: 0.0281). The regression intercept was 1.031, indicating little influence of population stratification or relatedness^52^. These findings suggest a strong genetic basis for TBS, which is pivotal for understanding bone integrity.

**Figure 1.**
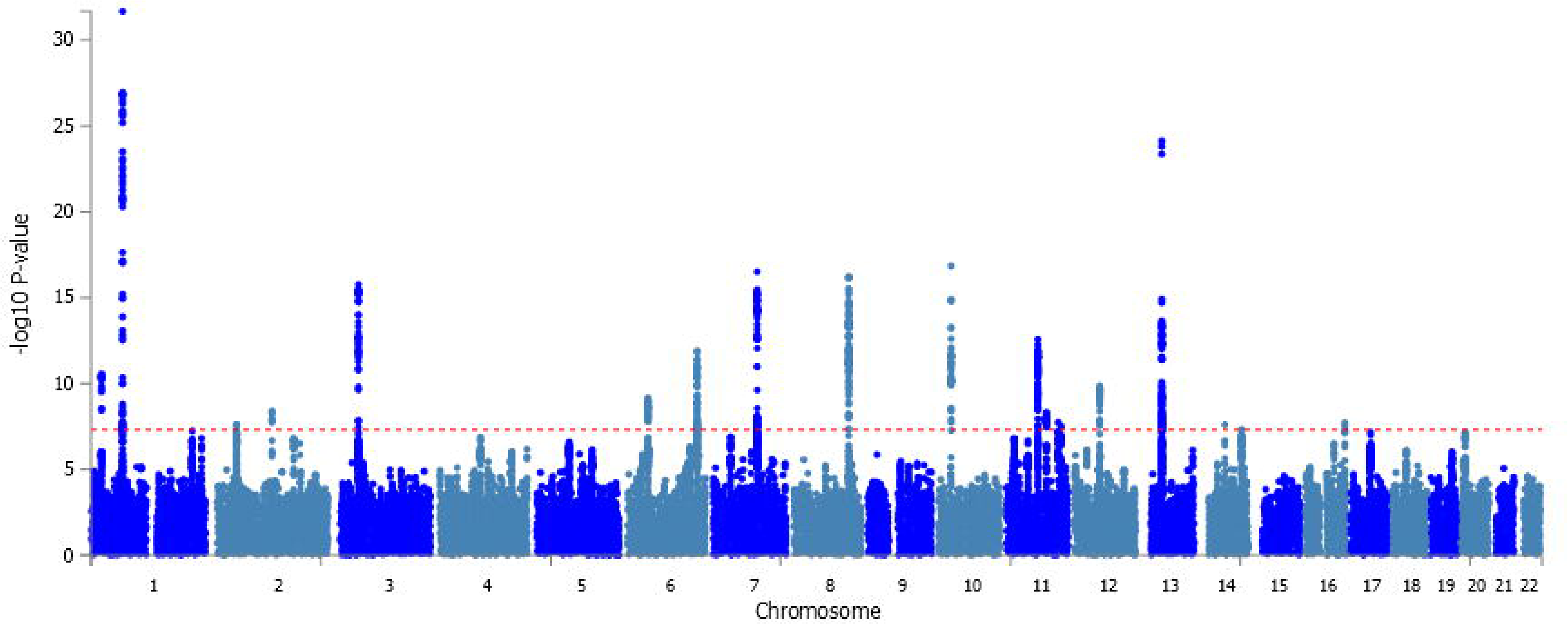
Genome-wide association study (GWAS) of trabecular bone score (TBS). (a) Manhattan plot of the GWAS on TBS. The P-values (−log10) are plotted against their respective positions on each chromosome. SNPs with P < 1×10^-5^ are shown. The red dotted line represents a significant threshold for the whole genome (P < 5×10^-8^). (b) Q–Q plot of the GWAS on TBS. SNPs with P-values < 1×10^-5^ are shown, and overlapping data points were excluded.

### 3.3 Functional mapping and annotation

Having identified key genetic variants associated with TBS, we next explored the effects of these variants on bone microarchitecture. Gene-based association analysis using MAGMA covered 19,002 protein-coding genes. Twenty-seven genes reached the genome-wide significance threshold (P < 2.63×10^-6^, 0.05/19,002; Fig. S1, Table S8). The *WLS* gene had the strongest association, with a P-value of 7.1962×10^-20^. Gene set analysis identified two gene sets significantly associated with TBS (P < 5× 10^-6^): “*RUNX1* regulates transcription of genes involved in Wnt signaling” (P = 1.4× 10^-6^), and “odontogenesis of dentin-containing tooth” (P = 2.27×10^-6^) (Table S9). The association of gene expression patterns in different tissues with bone microstructure deterioration was assessed by analyzing gene expression in 30 general tissues and 53 specific tissues from the GTEx v8 database. We found that gene expression in these tissue types was not significantly associated with bone microarchitecture (P < 0.001) (Table S10). These findings reveal mechanisms potentially governed by TBS-associated SNPs and set the stage for exploring the effects of these SNPs on health traits.

### 3.4 Association lookups with health traits

With a clearer understanding of the functional roles of these SNPs, we analyzed the associations of genetic factors with health traits by cross-referencing the identified SNPs with entries in the NHGRI-EBI GWAS Catalog^53^, the most comprehensive database of its kind. TBS-associated SNPs were correlated with several traits and diseases, including bone metabolism, bone integrity, and pathological disorders (inflammatory, autoimmune, cardiovascular, metabolic, and neuropsychiatric) (Table S7). These associations demonstrate the wide-ranging impact of bone health on overall health, indicating potential targets for therapeutic intervention and risk assessment.

Building on these systemic implications, we next focused on the impact of TBS-associated SNPs on BMD and fracture, which are crucial factors in musculoskeletal health. First, we conducted an in-depth analysis of TBS-associated loci using the largest GWAS dataset for BMD and fractures (Table S7). Among 19 TBS-associated loci, 11 had been identified in a study on areal BMD (aBMD)^9^ (five of 19 loci were associated with femoral neck BMD, and seven were associated with lumbar spine BMD), and locus rs3020332 was associated with fractures^54^ in individuals older than 18 years with medically, radiologically, or questionnaire-confirmed fracture at any skeletal site (Figure 2). To identify loci shared by bone microstructure, BMD, and fractures in the same UK Biobank cohort, we performed a GWAS of aBMD and fractures from UK Biobank data using the analytical criteria described above. Except for rs2143956, most TBS loci were associated with aBMD (segments L1 to L4, 13/19; heel, 16/19; spine, 12/19; total body, 12/19), and rs634277 was associated with fracture (fractured or broken bones in last 5 years). The identification of 18 loci shared by TBS and BMD and two loci linked to fracture risk indicates that bone strength and fractures share genetic components. Additionally, the discovery of a unique region (rs2143956, located at 14q22.2) associated with TBS highlights the distinct role of this region in bone microarchitecture, offering potential new avenues for targeted interventions and risk assessment.

**Figure 2.**
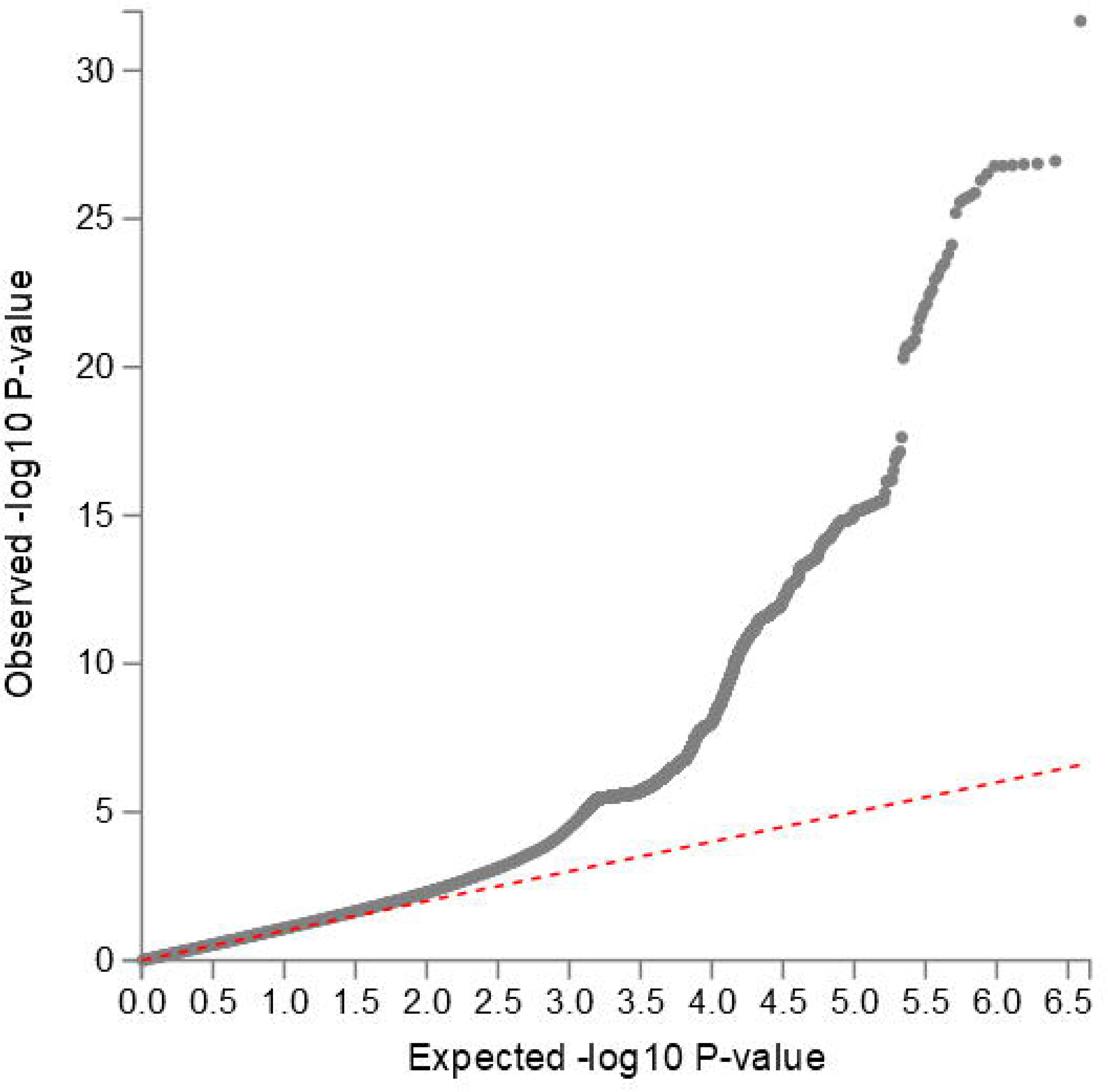
Associations of 19 significant genomic regions of TBS with bone mineral density (BMD) and fractures. The left heatmap shows the P-values of independent significant SNPs in a given genomic region in the GWAS of BMD and fracture types in the UK Biobank cohort. The right heatmap shows associations of independent significant variants from the GWAS analysis of BMD or fracture (P < 5 × 10^-8^ or P < 1 × 10^-5^) by the Genetic Factors for Osteoporosis Consortium (GEFOS).

### 3.5 Genetic correlation analysis

We performed a genetic correlation analysis to assess the genetic relationships of TBS with health traits. There were significant positive genetic correlations between TBS and lumbar spine BMD (r_g_ = 0.87, P = 2.68×10^-29^), femoral neck BMD (r_g_ = 0.74, P = 2.48×10^-24^), total body BMD (r_g_ = 0.81 for total body, P = 1.66×10^-58^), and grip strength (r_g_ = 0.16, P = 1.60×10^-5^) and a significant negative correlation between TBS and fracture risk (r_g_ = -0.47, P = 9.66×10^-13^). Moreover, TBS was suggestively associated with osteoarthritis (r_g_ = 0.12 for hip, P = 0.037; r_g_ = 0.10 for knee, P = 0.044), height (r_g_ = 0.08, P = 0.033), age at menarche (r_g_ = 0.14, P = 0.030), and type 2 diabetes (rg = 0.12, P = 0.016) (Fig. 3, Table S11).

**Figure 3.**
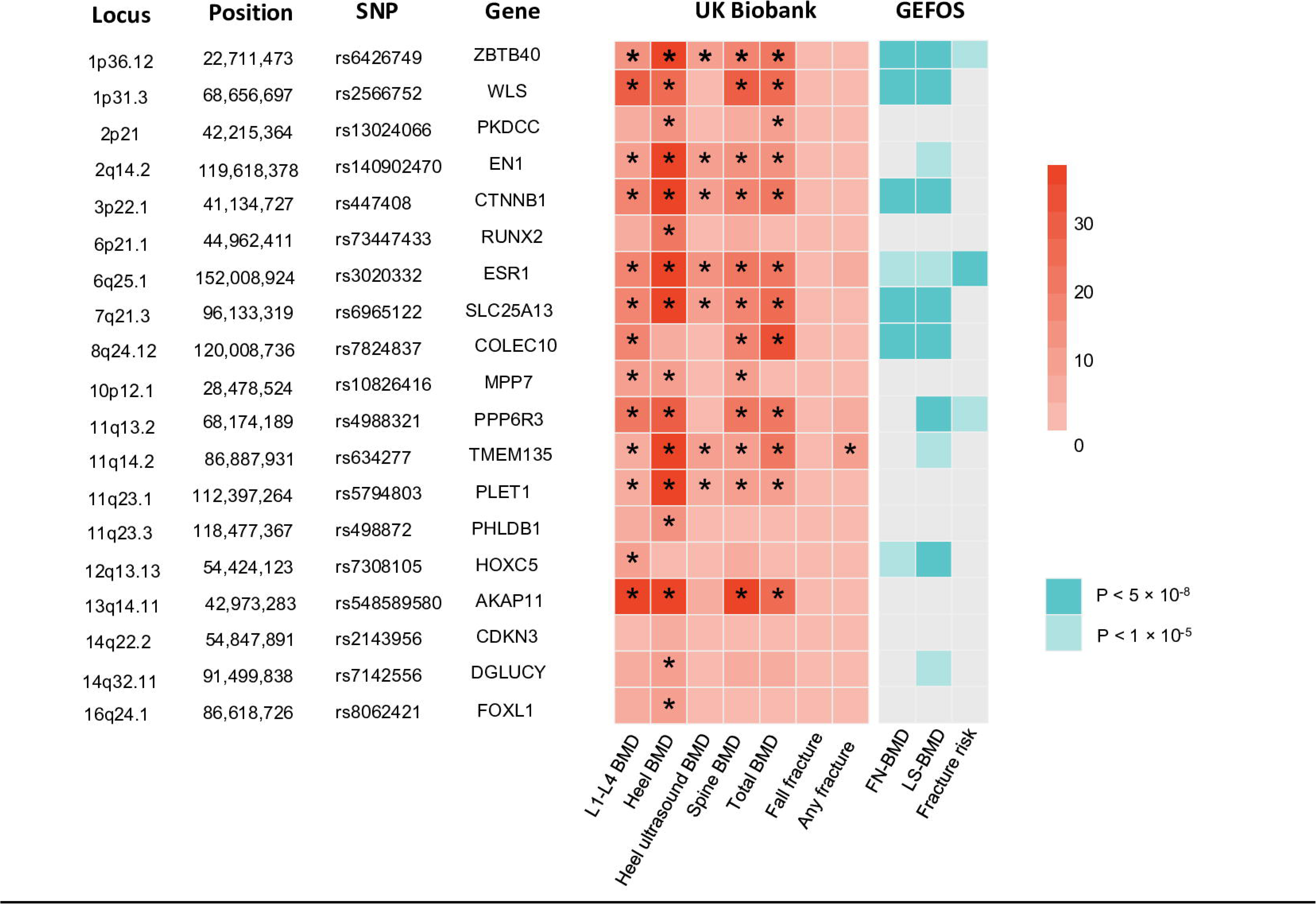
Genetic correlations of TBS with health traits and diseases. Genetic correlation (rG) and corresponding 95% confidence intervals (error bars) of TBS with health traits were estimated by linkage disequilibrium score regression. The size and direction of genetic correlations are indicated in different colors. Circles indicate a suggestive or significant correlation between two traits (p < 0.05), and diamonds indicate a lack of statistical significance.

### 3.6 Causal relationships

Based on the results of genetic correlation analysis and previous observational studies, we performed MR to assess causal relationships. MR uses genetic variants to infer causality between risk factors and health outcomes, thereby providing a more unbiased assessment than observational studies, which are susceptible to confounding.

Our MR analysis was designed to be bidirectional. First, overall health indicators and TBS were used as the exposure and outcomes, respectively, to assess the impact of overall health on bone microarchitecture (Fig. 4 and Table S12). After Bonferroni correction for multiple comparisons, genetically predicted RA was significantly negatively associated with TBS (β = -0.003, P = 1.14×10^-4^). In addition, the following conditions had a suggestive association with TBS: inflammatory bowel disease (β = -0.001, P = 0.030), low grip strength (β = -0.023, P = 0.005), cardiovascular diseases (β = -0.013, P = 0.031), asthma (β = -0.006, P = 0.032), fibroblastic disorders (β = 0.002, P = 0.018), and major depression (β = -0.013, P = 0.007).

**Figure 4.**
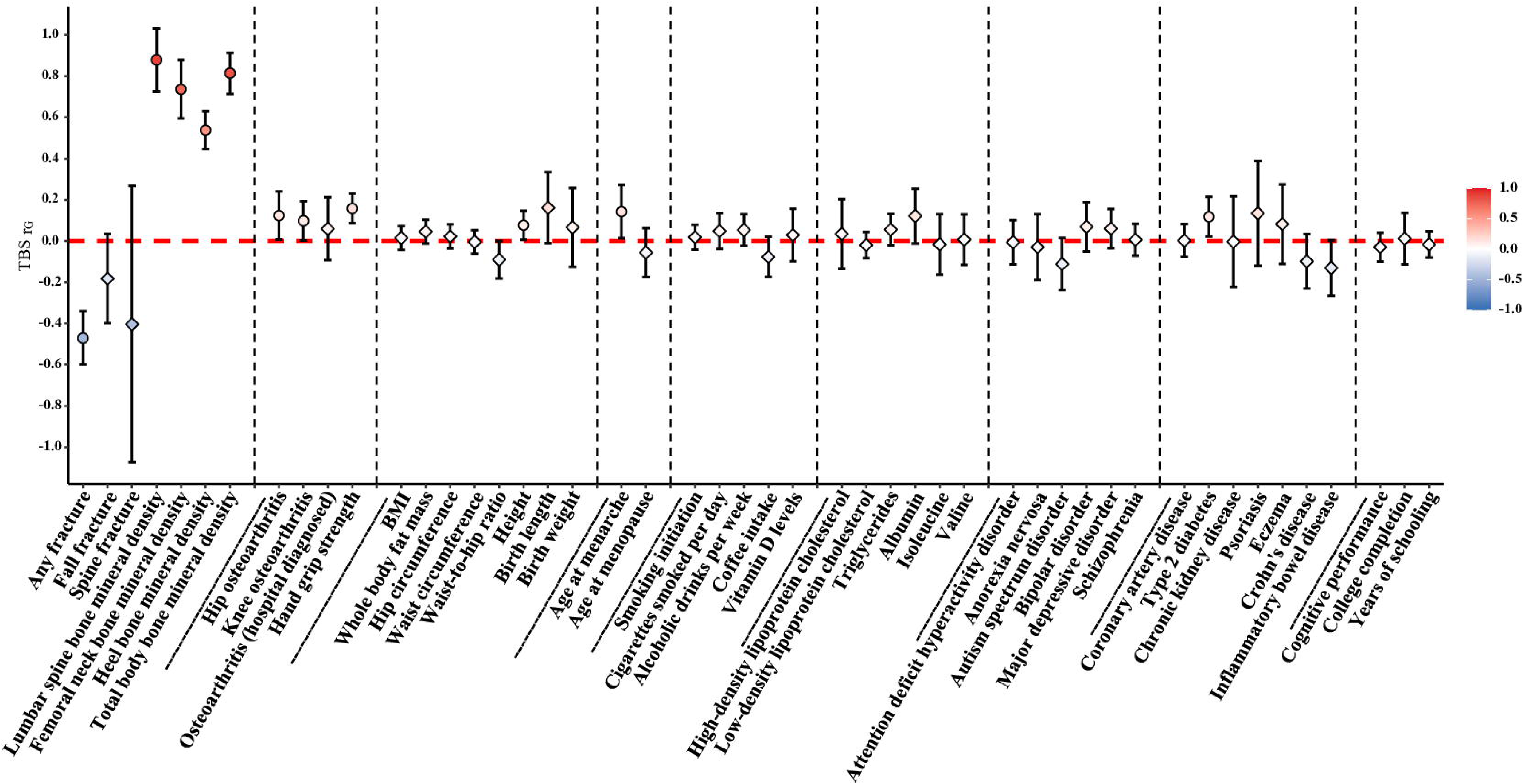
Mendelian randomization analysis of the causal effects of 17 clinical risk factors on trabecular bone score.

In the reverse-direction MR analysis, TBS and musculoskeletal disorders were considered the exposure and outcome, respectively (Fig. 5 and Table S13). After Bonferroni correction, genetically predicted TBS was significantly negatively associated with fracture risk (odds ratio [OR] = 0.003, P = 1.89 × 10^-8^) and suggestively associated with osteonecrosis (OR = 0.002, P = 0.040). In turn, TBS was not causally associated with ankylosing spondylitis (OR = 5.605, P = 0.579) and osteoarthritis of the hip and knee (OR = 2.169, P = 0.740).

**Figure 5.**
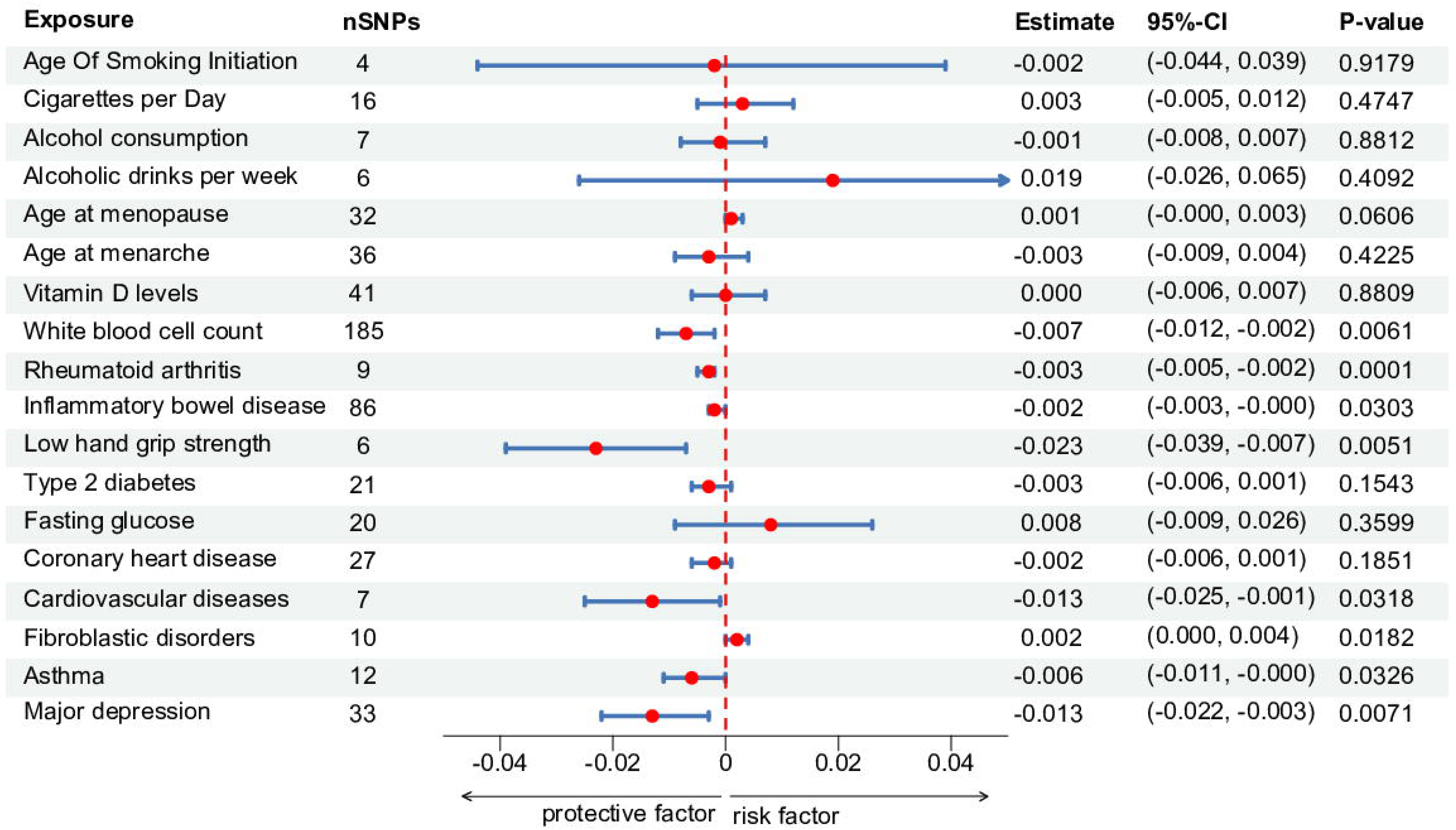
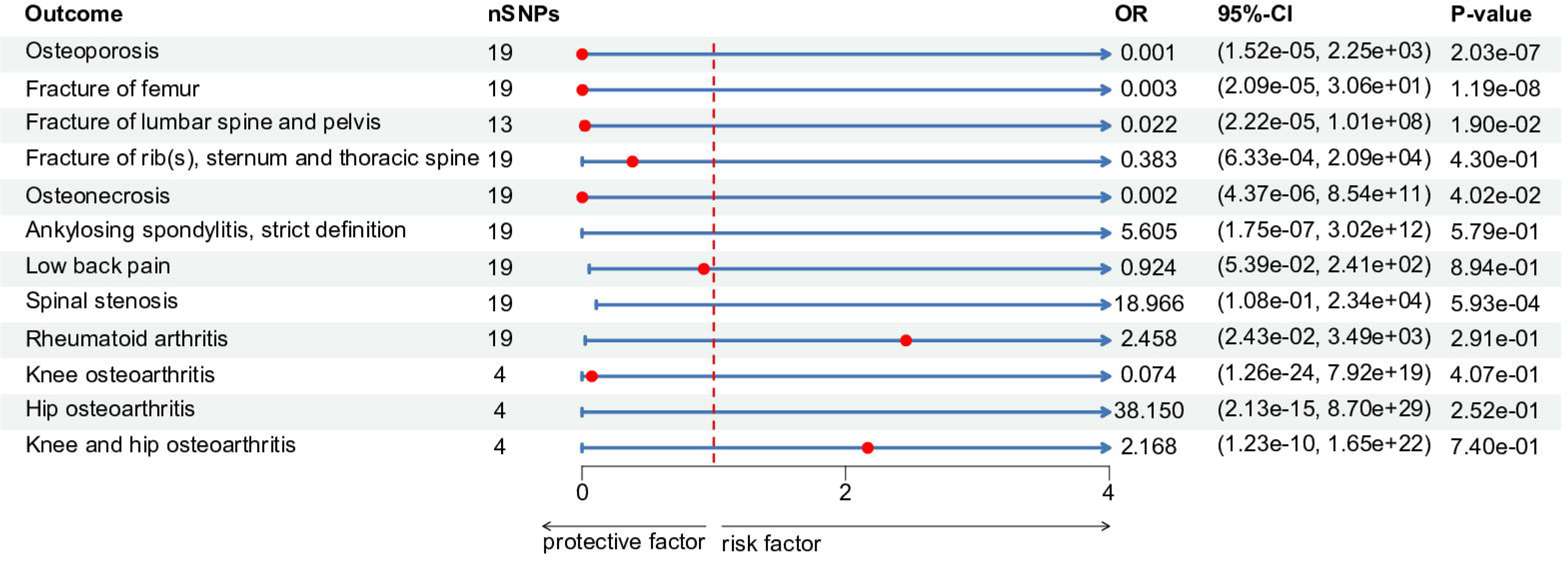
Mendelian randomization analysis of the causal effects of trabecular bone score on musculoskeletal disorders.

## 4. Discussion

This comprehensive genetic analysis of bone microarchitecture utilized TBS data from the UK Biobank as a surrogate phenotype and identified 19 loci associated with bone microarchitecture. These loci are shared by factors impacting bone integrity, including fractures, BMD, and grip strength. Furthermore, MR analysis indicated that RA was a significant risk factor for microarchitectural deterioration and that bone microarchitecture was strongly associated with osteoporosis, fractures, and spinal stenosis. Additionally, microarchitectural deterioration was suggestively associated with cardiovascular disease, inflammatory bowel disease, and depression, demonstrating the interplay between bone integrity and overall health.

GWAS analysis of TBS identified 75 significant SNPs across 19 loci. The most significant risk locus was located in chromosome 1p31.3, which contains the *WLS* gene, a crucial component of the Wnt signaling pathway involved in bone metabolism and remodeling.^55,56^. Additionally, TBS-associated genes were significantly enriched in the “*RUNX1* regulates transcription of genes involved in Wnt signaling” pathway, underscoring the importance of the Wnt pathway in bone microarchitecture. *RUNX1*, traditionally recognized for its role in hematopoiesis^57–59^, also contributes to skeletal development and bone homeostasis by regulating osteoblast and osteoclast activity^60–62^. *RUNX1* may regulate key genes in the Wnt pathway directly or indirectly, such as the expression of Wnt ligands, receptors, or downstream effectors. Wnt signaling may affect bone formation and remodeling by regulating the proliferation, differentiation, and activity of osteoblasts and osteoclasts. Furthermore, there is likely to be feedback regulation between *RUNX1* and the Wnt pathway, forming a sophisticated regulatory network to maintain the bone integrity. Further research is needed to elucidate the intricate interactions within this network and their implications for bone health.

TBS-associated SNPs were significantly associated with systemic diseases, particularly musculoskeletal disorders. The genetic linkage between TBS, osteoporosis, and fractures underscores the importance of assessing bone microarchitecture in clinical settings. Unique loci not associated with BMD, such as the T allele of rs2143956 in *CDKN3*, reveal novel aspects of bone microarchitecture that may be targeted for treating osteoporosis.

Our MR analysis evaluated the effects of RA on bone microarchitecture. Observational studies suggest that RA is associated with an increased risk of osteoporosis and fractures^63^. MR analysis demonstrated a significant causal effect of RA on microarchitectural deterioration, highlighting the negative impact of chronic inflammation on bone health. Inflammation is a critical mediator of osteoporosis risk in RA patients^64,65^. Pro-inflammatory cytokines contribute to joint destruction and bone loss. In addition, our findings suggest that inflammatory bowel disease is associated with TBS, although the association did not meet the stringent significance threshold after Bonferroni correction. The results also support the idea that chronic inflammation plays a role in bone health. The conflicting results on the causal association between RA and BMD at five sites^66,67^ suggest that the increased risk of osteoporosis and fractures in RA patients is more directly related to microarchitectural deterioration than to bone loss. Nonetheless, additional studies are needed to confirm this hypothesis.

Our findings indicate that TBS is a promising tool for assessing the risk of osteoporosis and fractures in RA patients, consistent with a previous study^68^ that showed that lumbar spine TBS had a higher ability than lumbar spine BMD to predict the presence of vertebral fractures in patients with RA.

Our genetic correlation and MR analyses reveal suggestive significant associations between bone microarchitecture and systemic diseases, including cardiovascular, psychiatric, and autoimmune diseases, suggesting that bone health is affected by overall health and pathological conditions. For instance, the causal relationship between TBS and cardiovascular health challenges the conventional understanding of risk factors for musculoskeletal diseases, suggesting that cardiovascular health metrics could serve as indicators of bone health.

This study has strengths. First, to our knowledge, this study is the most comprehensive evaluation of the genetic factors influencing bone microarchitecture to date, and represents the largest GWAS on TBS. The large sample size and extensive genetic data allowed for a robust analysis of the complex genetic architecture underlying bone microstructure. Second, leveraging the power of this large-scale GWAS, we are the first to assess the causal effect between bone microarchitecture and systemic health outcomes using a MR approach. This innovative application of genetic data provides novel insights into the potential causal relationships between bone health and various disease outcomes, opening new avenues for future research and clinical interventions.

This study has limitations. First, only participants aged 40-69 years were included in the analysis, thus limiting the phenotypic diversity of TBS. Second, other potential risk factors, such as BMI, were not included in the MR analyses. Third, in the two-sample MR analyses, we relied on GWAS data without access to individual-level data, preventing the accurate estimation of sample overlap and limiting the reliability of MR results, although we attempted to avoid sample overlaps by searching the relevant literature to make our selections. Fourth, while MR presupposes a linear relationship between risk factors and outcomes, the identification of marginal relationships, such as those between hypovitaminosis D and bone microarchitecture, does not negate the possibility of nonlinear relationships. Fifth, since we analyzed data from European populations, the results may not apply to other ethnic groups.

Future research can address the limitations of this study by (i) including participants from other age groups to enhance the generalizability of findings; (ii) evaluating other variables, including BMI, lifestyle, and nutrition, to unravel the relationships of genetic factors with bone microstructure; (iii) conducting cross-cultural studies to explore the impact of genetic backgrounds on bone health; (iv) utilizing advanced statistical models (e.g., nonlinear MR methods) to examine dynamic risk factor-outcome relationships; (v) integrating multi-omics data to reveal gene-environment-lifestyle interactions; and (vi) leveraging cutting-edge imaging technologies, such as high-resolution peripheral computed tomography, combined with machine learning to enable the early detection and individualized management of bone diseases.

In conclusion, our research examines the genetic and epidemiological facets of bone health, enriching the existing body of knowledge. We recommend incorporating TBS into clinical practice and elucidating the genetic complexities of bone microarchitecture and the influence of systemic diseases on bone quality. Our findings provide a foundation for understanding the intricate dynamics of bone health and devising targeted management strategies for musculoskeletal diseases.

## Data Availability

All data produced in the present study are available upon reasonable request to the authors

## Data sharing statement

The principal findings derived from the genome-wide association study (GWAS) and Mendelian randomization analyses are comprehensively presented within the main text and supplementary materials of this manuscript. For the Mendelian randomization analyses, we utilized GWAS data obtained from the IEU open GWAS database (https://gwas.mrcieu.ac.uk/). The complete GWAS summary statistics for trabecular bone score (TBS) are scheduled for public release in the immediate future. In the interim period, researchers interested in accessing these data may submit a request to the corresponding author. All reasonable requests will be considered, subject to any restrictions regarding participant privacy and data usage agreements.

## Author Contributions

CK-C, ZJW, and GS conceptualized and designed the study. GS conducted the experiments and performed data analysis, ZJP contributed to data preparation, cleaning, and analysis. GS drafted the initial manuscript. CK-C, ZJW, LH, and GN-L provided invaluable insights on the clinical interpretation of the findings and critically revised the manuscript. All authors have reviewed the paper, provided feedback, and approved the final version of the manuscript for submission.

## Acknowledgments

We thank the UK Biobank database and Dr. Celia Gregson from the University of Bristol for providing the trabecular bone score data for the UK Biobank. We also thank to Dr. Weichen Song from the School of Biomedical Engineering at Shanghai Jiao Tong University for his guidance on research methodology. Additionally, we acknowledge TopEdit LLC for the linguistic editing and proofreading during the preparation of this manuscript.

---

**Figure S1.**
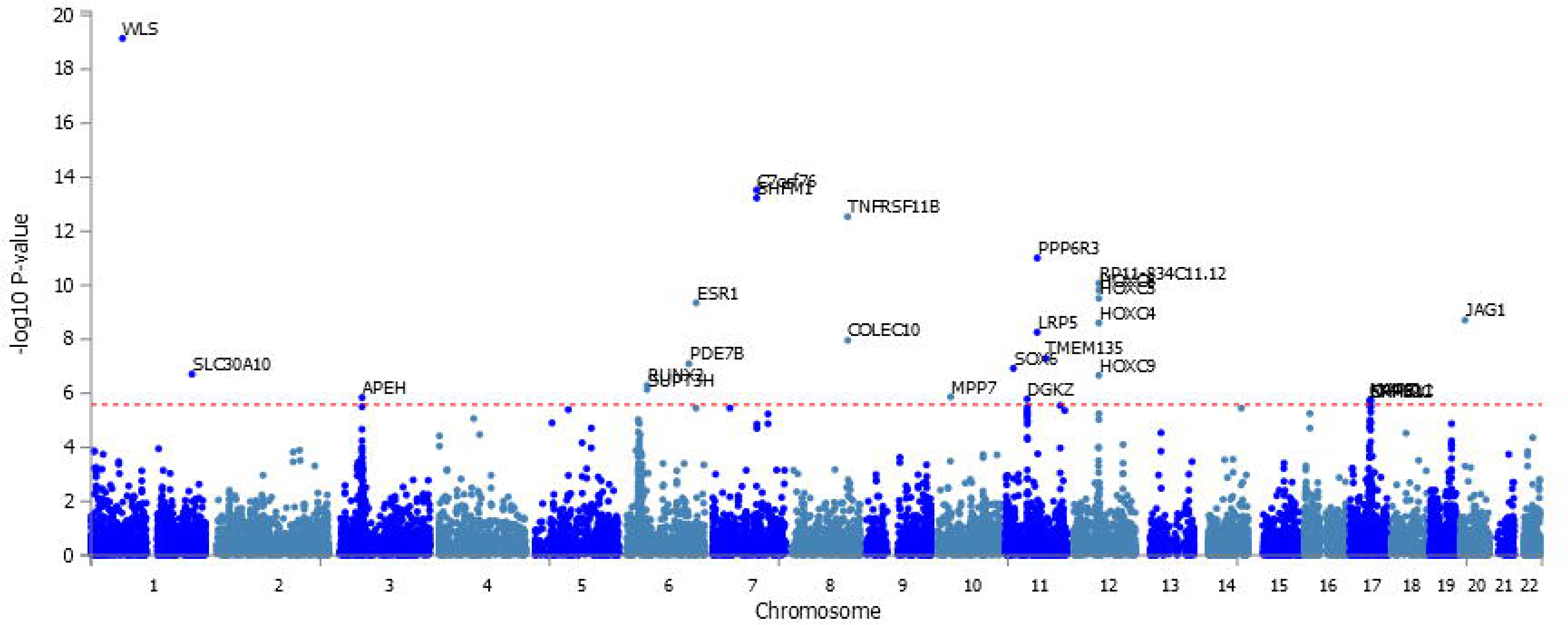
Manhattan plot of the gene-based analysis of trabecular bone score (TBS). The red line indicates genome-wide significance (p < 2.63× 10^-6^).

